# COVID-19 Booster Doses Reduce Sex Disparities in Antibody Responses among Nursing Home Residents

**DOI:** 10.1101/2024.10.23.24315989

**Authors:** Oladayo A. Oyebanji, Anna Yin, Nicholas Sundheimer, Vaishnavi Ragavapuram, Patrick Shea, Yi Cao, Philip A. Chan, Aman Nanda, Rohit Tyagi, Sakeena Raza, Nadia Mujahid, Yasin Abul, Alejandro B. Balazs, Jürgen Bosch, Christopher L. King, Sabra L. Klein, Stefan Gravenstein, David H. Canaday, Brigid M. Wilson

**Author notes:** **Corresponding authors:** Oladayo A. Oyebanji, David H. Canaday, Division of Infectious Diseases, Case Western Reserve University, 10900 Euclid Ave, BRB 1001, Cleveland, Ohio 44106-4984.

## Abstract

**Background:** Data suggests that antibody responses following COVID-19 vaccines are a correlate of protection. Some studies, including the clinical trials of COVID-19 mRNA vaccines, did not stratify and evaluate whether antibody responses to COVID-19 vaccines differed between the sexes or with aging. This gap in research is particularly relevant for older populations such as nursing home residents (NHR). We hypothesized that sex differences in vaccine-induced antibody responses may intersect with age and be diminished among older adults residing in nursing homes.

**Methods:** We analyzed serum samples from 638 NHRs collected serially after the primary two-dose series and three subsequent booster doses of mRNA SARS-CoV-2 vaccinations. We analyzed anti-Spike IgG and neutralizing antibody titers to the Wuhan and Omicron BA.4/5 variant strains. Mixed-effects models predicting log-transformed titers were estimated to compare responses across vaccine doses, focusing on sex-differential responses. For detected post-dose sex differences, additional sample times were analyzed to assess the duration of the difference.

**Results:** Following the primary series, female NHRs with a prior history of SARS-CoV-2 infection had significantly higher Wuhan anti-Spike antibodies and neutralizing antibody titers than male NHRs with differences persisting up to nine months post-vaccination. Subsequent monovalent booster doses and a bivalent booster dose eliminated this disparity. We did not detect any differential response to the Omicron BA.4/5 variant.

**Conclusions:** The blunting of sex differences in antibody response observed following the primary series by the 1st booster dose underscores the importance of booster vaccination in this population.

## BACKGROUND

Coronavirus disease 2019 (COVID-19) vaccines have played a vital role in mitigating the global impact of the pandemic. Examining the intricacies of vaccine efficacy and safety, an increasingly important factor has come to light -the role of biological sex differences in the immunological response to these vaccines, particularly among vulnerable populations such as nursing home residents (NHRs). Historically, there has been an acknowledgment of the substantial influence of sex differences on immune responses to various infections and vaccines (1-4). The emergence of the COVID-19 pandemic and the subsequent rollout of vaccines have provided a unique opportunity to explore these disparities in greater detail, often overlooked in clinical trials (5-7).

Previous studies suggest that sex-based differences in the immune response to COVID-19 may significantly affect disease outcomes (8-10). However, we lack comprehensive studies focusing on this topic within the specific context of NHRs. Generally, females exhibit heightened inflammatory, antiviral, and humoral immune responses compared to males, with roles for genes and sex steroid hormones, like estradiol (11,12). Similarly, the immune response in older persons, especially females, shows a progressive decline, highlighting the intricate interaction of sex and age in immune function (13-15). This decline in immune function with aging, known as immunosenescence, particularly affects the efficacy of vaccines and increases susceptibility to infections and poorer health outcomes in older populations. Furthermore, sex differences in COVID-19 outcomes, with males exhibiting more severe illness, have prompted investigation into differential immune responses (16-18). These findings collectively emphasize the need for a nuanced understanding of sex-specific immunological responses to COVID-19 vaccines among distinct populations such as institutionalized older adults.

In a previous study involving a cohort of NHRs and healthcare workers, we showed that female NHRs elicited a higher T-cell response than male NHRs following repeated mRNA vaccinations (19). This current study investigates whether there are variations in humoral immune responses to COVID-19 vaccines between male and female NHRs. We aim to contribute to the growing body of knowledge surrounding biological sex differences in humoral responses to COVID-19 vaccines, specifically focusing on NHRs, who are susceptible to severe COVID-19 outcomes due to their advanced age and underlying health conditions.

## METHODS

### Ethical approval

This study was approved by the Western-Copernicus Group Institutional Review Board (WCG IRB) with the protocol number STUDY20211074. All participating residents or their legally authorized representatives provided informed consent to be enrolled. The study is in accordance with the 1964 Declaration of Helsinki and its later amendments.

### Study Design and Study Population

The current analysis is part of an ongoing study (20-24) in which NHRs are consented and serially sampled before and after each SARS-CoV-2 vaccine dose. Participants were recruited from 18 nursing homes in Ohio and 16 nursing homes in Rhode Island. Comorbidity and functional status were added to the study data collection, based on chart review of subject health records, after enrollment began. Thus, these variables were collected for most, but not all, of the subjects. Residents who received SARS-CoV-2 mRNA vaccines [(BNT162b2 (Pfizer-BioNTech) or mRNA-1273 (Moderna)] were included, and those who received other vaccines were excluded. Participants typically received their first monovalent booster dose 8-9 months after the primary vaccination series, and their second monovalent booster 4 to 6 months after the first booster. In this current study, we report results from blood samples obtained at time points following vaccination: approximately 14 days, 4-6 months, and 8-9 months post-primary vaccination series; and 14 days post-first, second monovalent booster, and post-bivalent booster (Fig. 1). The primary vaccination series and the first and second monovalent boosters were Wuhan-based mRNA vaccines while the bivalent booster consisted of Wuhan and Omicron BA.4/5 strains. All samples were collected between December 2020 and December 2022. In the setting of breakthrough infection during the study, the subject’s samples collected from the breakthrough through the next vaccine dose were excluded from this analysis.

**Fig. 1:**
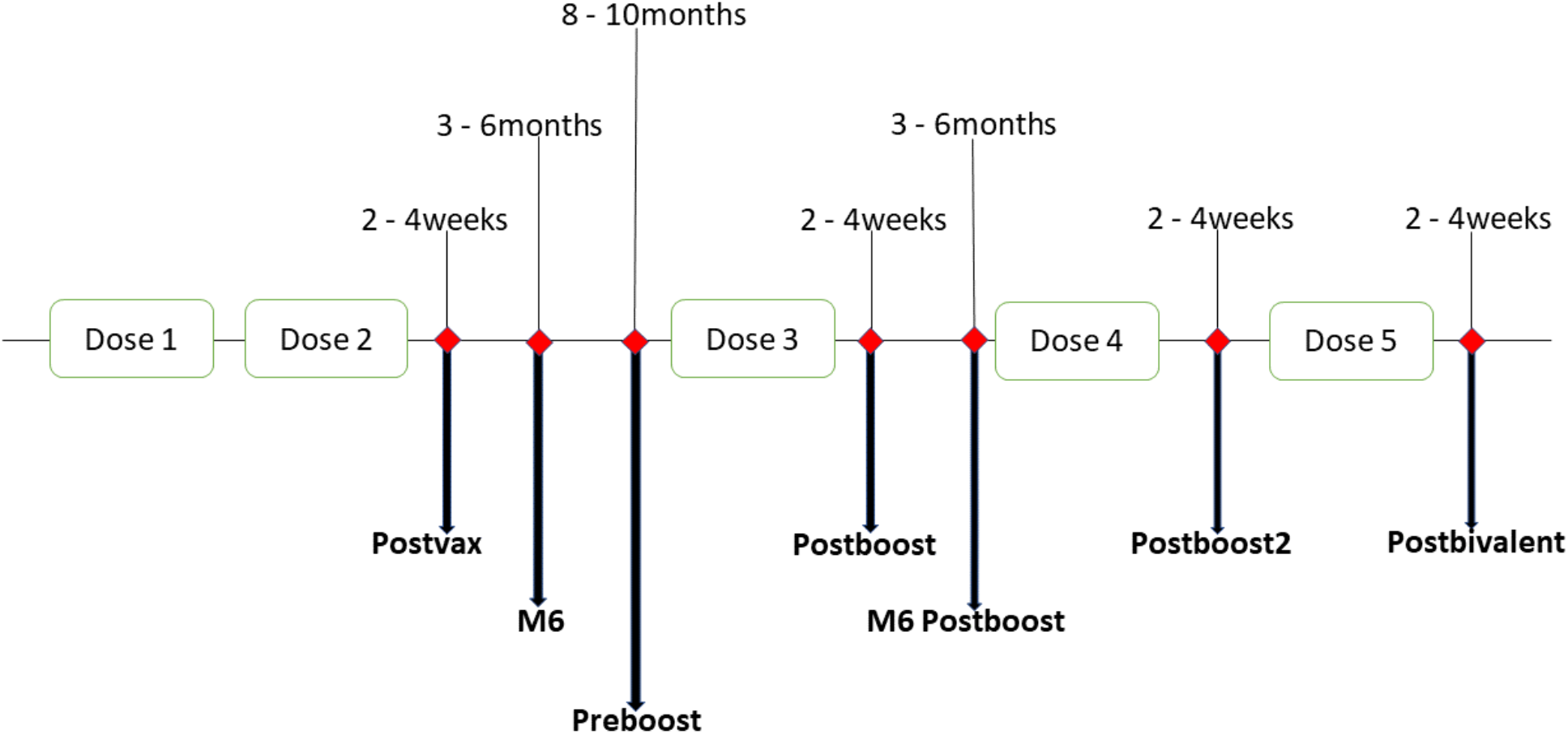
Timeline of blood sampling from participants. Serum samples were collected from participants at different time points after mRNA vaccination. The big diamond represents sampling times, generally 2-4 weeks and 3-6 months after vaccination. Doses 1 and 2 are the 1st and 2nd doses, respectively, given 3 weeks apart. Doses 3 and 4 are the 1st and 2nd Wuhan-based monovalent booster doses, given at least 6 months after the previous dose. Dose 5 is a Wuhan-Omicron BA.4/5-containing bivalent booster dose. While many of our participants did not receive the 2nd monovalent booster (Dose 4), those who did, got the bivalent booster within 4-6 months.

Participants were deemed “infection prior” if they had a prior SARS-CoV-2 infection at the time of each sampling based on: 1. Prior documentation in their medical chart of a positive polymerase chain reaction (PCR) or antigen test; or 2. An increase in SARS-CoV-2 antibody levels beyond variation of the assay, that could not be explained by vaccination e.g. rise in Spike-specific and N-antigen-specific antibodies.

### Anti-Spike Assay

We assessed vaccine-induced antibody response using bead-multiplex immunoassay for anti-spike for SARS-CoV-2 wild-type (Wuhan-Hu-1) strain and BA.4/5 variants as previously described (20). Stabilized full-length spike protein (aa 16-1230, with furin site mutated and recombinant SARS-CoV-2 S(1-1208)-2P-3C-His8-TwinStrep) from Wuhan and SARS-CoV-2 S-2P(1-1208)-3C-His8-TwinStrep BA.5 from Omicron BA.4/5 variants and full-length N (aa1-419) from Wuhan, obtained from the Frederick National Laboratory (FNL) were conjugated to magnetic microbeads (Luminex) and Magpix assay system (BioRad, Inc). Anti-Wuhan spike IgG levels were in Binding Antibody Units (BAU)/mL based on the FNL standard, and anti-spike BA.4/5 are shown in arbitrary units (AU)/mL.

### SARS-CoV-2 Pseudovirus Neutralization Assay

We produced lentiviral particles pseudotyped with spike protein based on the Wuhan and BA.4/5 strains as previously described to define the neutralizing activity of vaccine recipients’ sera against coronaviruses (25). We performed three-fold serial dilutions that ranged from 1:12 to 1:8748 and added 50–250 infectious units of pseudovirus for 1 hour. 50% pseudoviral neutralizing antibody titers (pNT50) values were calculated by taking the inverse of the 50% inhibitory concentration value for all samples with a pseudovirus neutralization value of 80% or higher at the highest serum concentration. The lower limit of detection (LLD) of this assay is 1:12 dilution.

### Statistical analysis

Separate models predicting vaccine response by dose, sex, prior infection, and all interactions of these 3 variables were estimated for each combination of strain (Wuhan and BA.4/5) and assay (anti-spike antibodies and neutralizing titers). As some subjects were sampled repeatedly, mixed-effects linear regression models predicting log-transformed titers were estimated to adjust for correlated outcomes within subjects using random intercepts. Model assumptions were checked and marginal mean sex differences were tested using model contrasts for each dose combination and prior SARS-CoV-2 infection.

For doses with detected sex differences post-vaccination, we analyzed additional samples from the post-vaccination subjects obtained before the next dose to test if the observed sex differences persisted over time using the modeling approach described above. Samples obtained 150-210 days and 240-300 days post-vaccine were grouped as 6-month and 9-month post-dose samples, respectively.

For those subjects with infections before the post-primary series sample, we compared the available dates of prior infections between males and females using a Wilcoxon rank sum test. To assess for possible sex differences in attrition over time due to death, we identified subjects with a study withdrawal due to death in the year following primary series vaccination and compared death rates by sex using a Fisher’s exact test.

Results were considered statistically significant at a two-sided alpha of 0.05. All analyses were performed in R version 4.2.2 using nlme and emmeans packages for model and contrast estimation.

## RESULTS

Our study involved 638 NHRs in total throughout the study and had 60 or more NHRs of each sex for each of the 4 vaccine doses. Across all time points, the participants are predominantly of white ethnicity. Female NHRs had a median age of 76 to 83 and were significantly older than the male NHRs, with a median age range of 73 and 75 (Table 1). In addition to advanced age, this cohort had a high burden of comorbidities and reduced functional status (Supplemental Table 1). Comorbidities and functional status were not collected from some of the earliest enrolled subjects in our study; thus, this data is missing for 29% of the post-primary series cohort. Regarding functional status, many more male NHRs were completely independent. Among those in the cohort with available comorbidity data, we observed higher rates of COPD and heart failure among males. Female NHRs had more dementia while the rate of diabetes mellitus and immunosuppressive illnesses or immunomodulatory medications were similar for both sexes. After the primary vaccination series, similar rates of SARS-CoV-2 infection were observed across both sexes at each vaccine dose (Table 1).

**Table I:** Demographics of Nursing Home Residents (NHRs) by vaccine dose.

|  |  | **Post-Primary**<br>**Series** | **Post-1st**<br>**Monovalent**<br>**Booster** | **Post-2nd**<br>**Monovalent**<br>**Booster** | **Post-Bivalent**<br>**Booster** |
| --- | --- | --- | --- | --- | --- |
| Female | N subjects | 60 | 157 | 120 | 133 |
|  | Age | 83 (73, 88.5) | 78 (70, 86) | 78.5 (69.8, 88) | 76 (68, 87) |
|  | Race/Ethnicity:<br>White Non-Hispanic | 52 (87%) | 127 (81%) | 97 (81%) | 106 (80%) |
|  | Race/Ethnicity:<br>Black Non-Hispanic | 6 (10%) | 28 (18%) | 20 (17%) | 26 (20%) |
|  | Race/Ethnicity:<br>Hispanic | 1 (2%) | 1 (1%) | 2 (2%) | 0 (0%) |
|  | Race/Ethnicity:<br>Other/Missing | 1 (2%) | 1 (1%) | 1 (1%) | 1 (1%) |
|  | Prior infection | 28 (47%) | 81 (52%) | 83 (69%) | 101 (76%) |
| Male | N subjects | 92 | 224 | 81 | 128 |
|  | Age | 75 (70, 83) | 74 (68, 83) | 74 (66, 81) | 73 (66, 78.25) |
|  | Race/Ethnicity:<br>White Non-Hispanic | 79 (86%) | 177 (79%) | 60 (74%) | 92 (72%) |
|  | Race/Ethnicity:<br>Black Non-Hispanic | 13 (14%) | 39 (17%) | 17 (21%) | 31 (24%) |
|  | Race/Ethnicity:<br>Hispanic | 0 (0%) | 4 (2%) | 2 (2%) | 2 (2%) |
|  | Race/Ethnicity:<br>Other/Missing | 0 (0%) | 4 (2%) | 2 (2%) | 3 (2%) |
|  | Prior infection | 41 (45%) | 113 (50%) | 49 (60%) | 92 (72%) |

We detected sex differences in antibody responses following the primary vaccination series among subjects with prior SARS-CoV-2 infection such that the observed geometric mean titer (GMT) of Wuhan anti-Spike antibodies in females was 3.2 times that of males (2431 vs. 755, p = 0.007) and the GMT of Wuhan neutralizing titers was 2.8 times that of males (1742 vs. 623, p= 0.004) (Table 2). These sex differences, undetected before initial vaccination, persisted and were statistically significant when examined in a linear mixed-effects model, adjusted for repeated samples within subjects across vaccine doses. When focusing on the post-vaccination time points before the first monovalent booster in residents with post-primary series data (n = 152) using similar models, we found that the sex-based difference in immunological response among prior-infected subjects persisted at 6 months (model estimated geometric mean titer rise, GMTR = 4.2, p = 0.003) and 9 months for the spike protein (model estimated GMTR = 16.3, p < 0.001) and at 6 months for neutralization titers (model estimated GMTR = 2.8, p = 0.014) (Figure 2). Among prior-infected NHRs sampled post-primary series, there was no statistically significant difference in the time elapsed since prior COVID-19 infection between males and females with medians of 87 days and 84 days, respectively (Wilcoxon p = 0.55). Among 4 mortality events in this post-primary series cohort observed in the year following primary series vaccination, 3 were men.

**Table II:** GMT by assay, vaccine dose, prior infection status for male & female nursing home residents with model-estimated ratio female to male and model p-values.

| Vaccine dose | Strain | Status | Assay | GMT (CI), Female | GMT (CI), Male | Crude Ratio, F/M | Adjusted Ratio, F/M | pval |
| --- | --- | --- | --- | --- | --- | --- | --- | --- |
| Post-Primary Series | Wuhan | Naive | Neut | 85<br>(50,143) | 108<br>(74,158) | 0.79 | 0.96<br>(0.53,1.75) | 0.892 |
|  |  |  | Anti-Spike | 131 (62,275) | 234<br>(145,378) | 0.56 | 0.67<br>(0.35,1.29) | 0.227 |
|  |  | Prior | Neut | 1742<br>(844,3595) | 623<br>(319,1218) | **2.8** | **2.61**<br>**(1.35,5.02)** | **0.004** |
|  |  |  | Anti-Spike | 2431<br>(1212,4875) | 755<br>(403,1413) | **3.22** | **2.83**<br>**(1.34,5.99)** | **0.007** |
| Post-1st Monovalent | Wuhan | Naive | Neut | 381<br>(207,701) | 433<br>(314,596) | 0.88 | 0.97<br>(0.56, 1.65) | 0.897 |
|  |  |  | Anti-Spike | 1745<br>(1115,2731) | 2055<br>(1414,2986) | 0.85 | 0.87<br>(0.55,1.37) | 0.551 |
|  |  | Prior | Neut | 1336<br>(786,2271) | 995<br>(666,1487) | 1.34 | 1.36<br>(0.77,2.41) | 0.291 |
|  |  |  | Anti-Spike | 6521<br>(4772,8912) | 7521<br>(5786,9774) | 0.87 | 0.83<br>(0.54,1.29) | 0.408 |
| Post-2nd Monovalent | Wuhan | Naive | Neut | 891<br>(503,1579) | 827<br>(484,1413) | 1.08 | 0.9<br>(0.45,1.79) | 0.76 |
|  |  |  | Anti-Spike | 1642<br>(870,3098) | 1571<br>(877,2815) | 1.05 | 0.93<br>(0.44,1.97) | 0.851 |
|  |  | Prior | Neut | 1427<br>(1043,1952) | 1271<br>(856,1886) | 1.12 | 1.25<br>(0.66,2.35) | 0.489 |
|  |  |  | Anti-Spike | 3979<br>(3018,5246) | 2769<br>(1914,4006) | 1.44 | 1.3<br>(0.77,2.2) | 0.326 |
|  | Omicron BA.4/5 | Naive | Neut | 232<br>(113,475) | 177<br>(89,353) | 1.31 | 1.14<br>(0.51,2.54) | 0.742 |
|  |  |  | Anti-Spike | 990<br>(562,1743) | 1333<br>(888,2000) | 0.74 | 0.68<br>(0.38, 1.2) | 0.181 |
|  |  | Prior | Neut | 1074<br>(697,1654) | 957<br>(498,1840) | 1.12 | 1.34<br>(0.66,2.71) | 0.41 |
|  |  |  | Anti-Spike | 2707<br>(1958,3744) | 2247<br>(1705,2961) | 1.2 | 1.05<br>(0.69,1.58) | 0.828 |
| Post-Bivalent | Wuhan | Naive | Neut | 1532<br>(884,2656) | 2015<br>(1305,3111) | 0.76 | 0.89<br>(0.4,1.97) | 0.774 |
|  |  |  | Anti-Spike | 2605<br>(1556,4360) | 3630<br>(2404,5480) | 0.72 | 0.72<br>(0.35,1.49) | 0.38 |
|  |  | Prior | Neut | 2975<br>(2118,4178) | 2277<br>(1371,3782) | 1.31 | 1.02<br>(0.58,1.81) | 0.937 |
|  |  |  | Anti-Spike | 3100<br>(2414,3980) | 3626<br>(2736,4807) | 0.85 | 0.92<br>(0.6,1.41) | 0.714 |
|  | Omicron<br>BA.4/5 | Naive | Neut | 1187<br>(558,2524) | 982<br>(539,1789) | 1.21 | 1.63<br>(0.66,3.98) | 0.279 |
|  |  |  | Anti-Spike | 1046<br>(707,1547) | 1354<br>(956,1917) | 0.77 | 0.71<br>(0.41,1.24) | 0.224 |
|  |  | Prior | Neut | 2434<br>(1709,3466) | 1630<br>(936,2840) | 1.49 | 1.44<br>(0.74,2.79) | 0.271 |
|  |  |  | Anti-Spike | 1361<br>(1104,1678) | 1522<br>(1165,1989) | 0.89 | 0.92<br>(0.66,1.28) | 0.622 |

Anti-Spike is measured in BAU/ml while Neutralization (Neut) is in pNT50. BAU/ml: Binding Antibody Unit per milliliter, pNT50: Pseudoneutralization titer 50, GMT: Geometric Mean Titer, CI: Confidence Interval, F: Female, M: Male

**Fig. 2:**
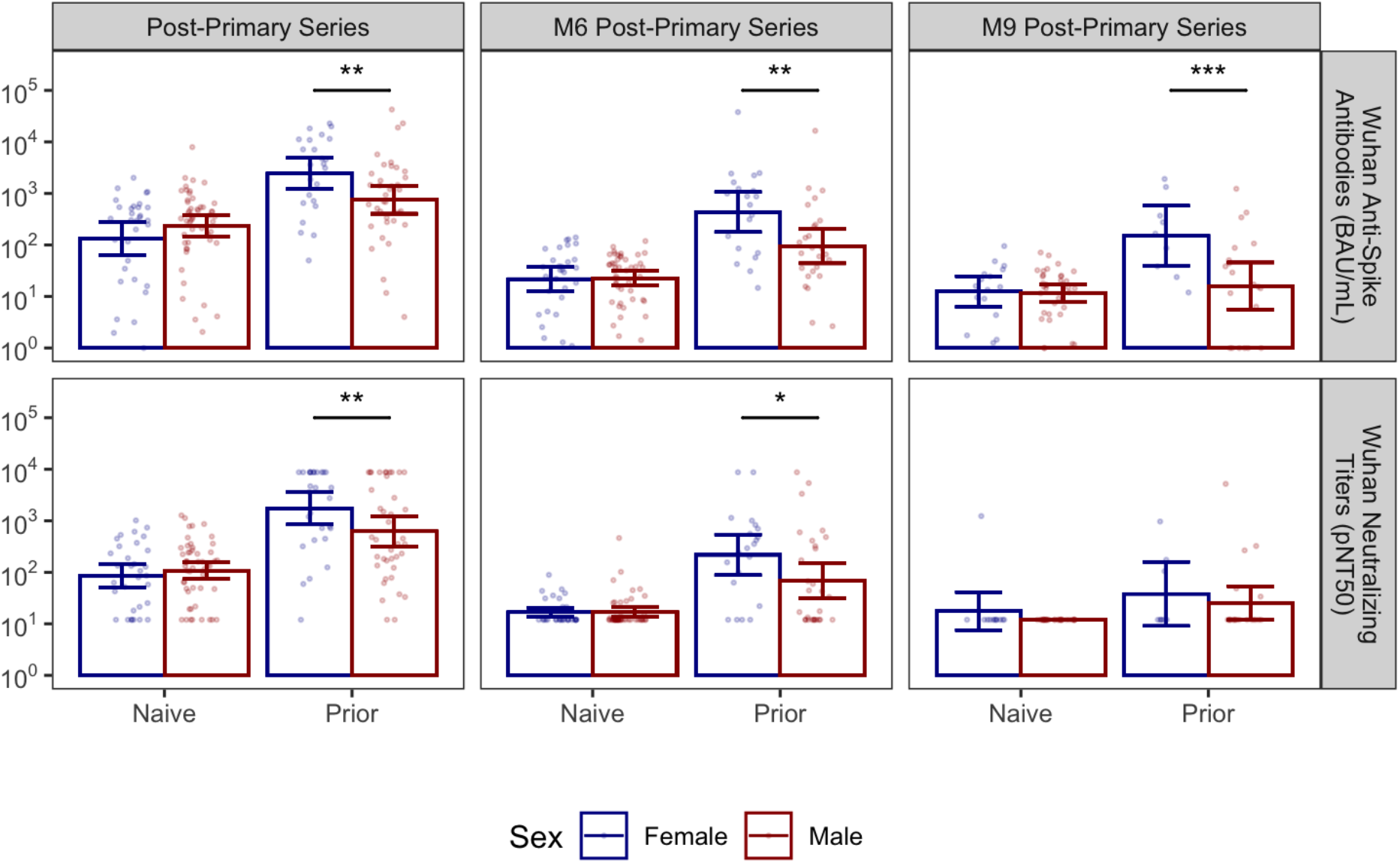
Anti-Wuhan Spike and neutralizing antibody titers over time among female and male NHRs. The bar graphs show the kinetics of anti-spike (upper panel) and neutralizing (lower panel) antibodies against the Wuhan strain across different time points among female and male NHR. Wuhan anti-Spike is measured in BAU/mL. The lower limit of detection of the neutralization assay was 1:12, while the upper limit was 1:8748. Post-primary series sera were taken 2-4 weeks after the 2nd vaccine dose, completing the primary series, while M6 and M9 post-primary series sera were taken 6-8 months and 7-10 months later, respectively. Bars and whiskers show GMT with 95% CI. Blue: Female, Red: Male. Naive subjects: no prior infection, Prior subjects: previously infected. Male and female subjects were compared with model contrasts after estimating mixed-effects linear models predicting log-transformed titer with the interaction of sex, prior infection, and sample time within strain and assay. ^*, **, ***^ are significance levels of model contrasts with p<0.05, <0.01, <0.001 respectively. BAU/ml: Binding Antibody Unit per milliliter, pNT50: Pseudoneutralization titer 50, NHR: Nursing Home Residents, GMT: Geometric Mean Titer, CI: Confidence Interval

We did not detect sex differences in vaccine response as measured by these assays to any of the three booster doses examined stratified by prior SARS-CoV-2 infection; nor did we detect sex differences in infection-naive subjects following the primary vaccination series (Figure 3A). We also did not detect sex differences when comparing Omicron BA.4/5 anti-Spike antibodies and neutralizing titers for the second monovalent booster and first bivalent booster doses (Figure 3B).

**Fig. 3:**
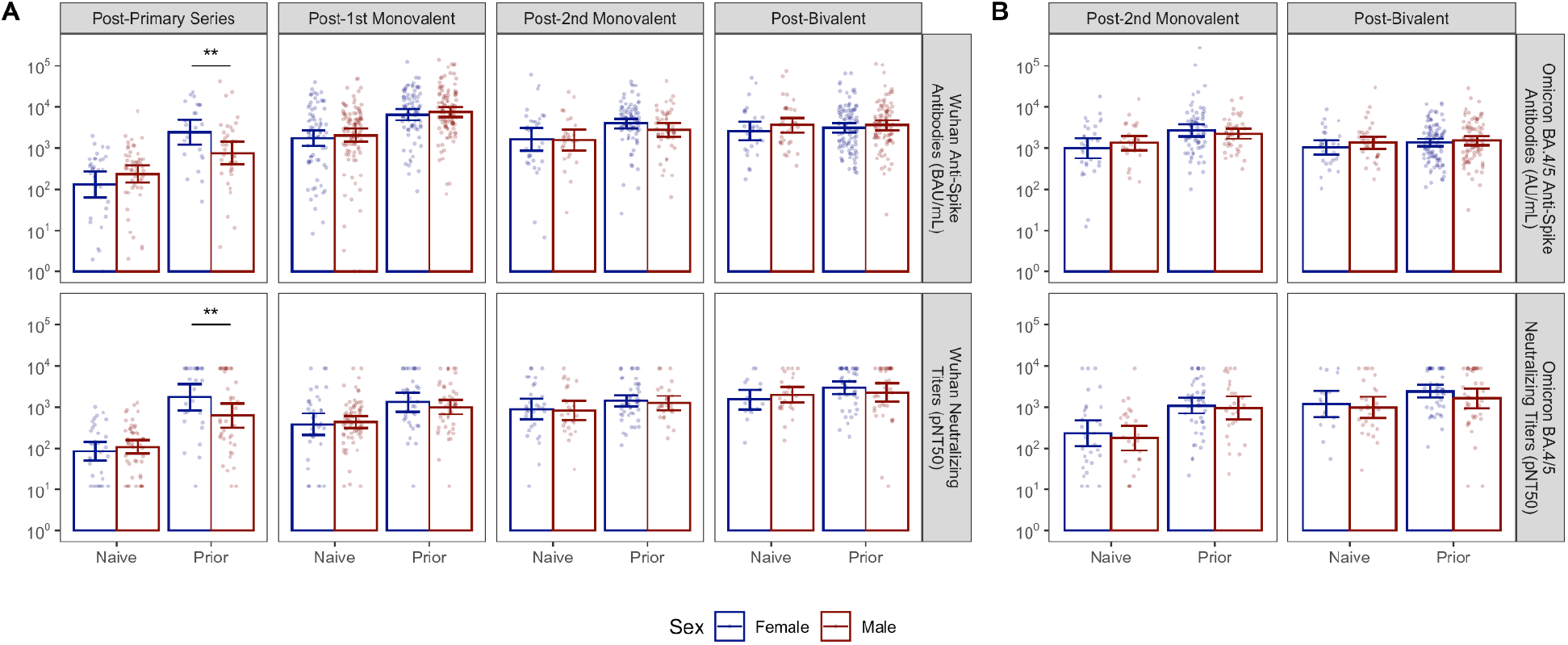
Anti-Spike and Neutralizing Antibody titers against Wuhan (Panel A) and Omicron BA.4/5 (Panel B) strains across booster doses among female and male NHR. The bar graph shows the post-vaccination anti-spike and neutralizing antibody titers against the Wuhan and Omicron strains across the boosters among female and male NHR. Wuhan anti-spike is measured in BAU/mL. The lower limit of detection of the neutralization assay was 1:12, while the upper limit was 1:8748. Post-vaccination sera were taken 2-4 weeks after each dose. Bars and whiskers show GMT with 95% CI. Blue: Female, Red: Male. Naive subjects: no prior infection, Prior subjects: previously infected. Male and female subjects were compared with model contrasts after estimating mixed-effects linear models predicting log-transformed titer with the interaction of sex, prior infection, and vaccine dose within strain and assay. ^*, **, ***^ are significance levels of model contrasts with p<0.05, <0.01, <0.001 respectively. BAU/ml: Binding Antibody Unit per milliliter, pNT50: Pseudoneutralization titer 50, NHR: Nursing Home Residents, GMT: Geometric Mean Titer, CI: Confidence Interval

## DISCUSSION

Understanding biological sex differences in the immune response to vaccination may help optimize vaccine efficacy and develop targeted interventions. Our study investigated the potential influence of biological sex on the humoral response to COVID-19 mRNA vaccines among NHRs.

Notably, we observed a significant sex-based disparity in antibody levels following the primary vaccination series among prior SARS-CoV-2-infected residents, with females exhibiting substantially higher levels of Wuhan anti-Spike antibodies and neutralizing titers compared to their male counterparts. While this aligns with some evidence highlighting sex-based differences in immune responses to viral infections and vaccinations (26-28), it remains unclear why this disparity was not present among the SARS-CoV-2 infection-naive residents in our study, as reported by Shapiro et al (29). This may have been influenced by factors such as different ages between naïve and prior residents, and comorbidities, among others. However, similar to our findings, in a large multicenter study of NHRs, Trevisan et al did not observe any sex differences in antibody response among naive residents who received 2 doses and prior-infected residents who received only 1 dose of the mRNA vaccines (30). In contrast to the vaccination regimen in that study, the prior-infected residents in our study received 2 doses of the mRNA vaccine. This additional antigenic exposure could have provided a window to enhance the disparity between the sexes in our study.

It is well established that females generally mount more robust and durable immune responses to infections and vaccinations, which may be due, in part, to the modulating influence of sex steroid hormones, particularly estradiol, on the immune system (31-33). Estrogen has been shown to enhance the production of antibodies and promote a robust immune response to viral infections, as its signaling pathways are intricately involved in the regulation of immune responses to viral infections (11,12,34,35). Androgenic hormones, such as testosterone, appear to have a dampening effect on immune functions (36). The heightened vaccine-induced antibody production among the prior-infected female residents in our study could be due to the dual antigenic exposure via infection and vaccination, resulting in a significant disparity between the sexes. It is essential to emphasize that while sex differences were observed following the primary series, the overall immunogenicity of the vaccines in both males and females is substantial, highlighting the importance of vaccination in this vulnerable population.

This disparate increase in antibody response among prior-infected female residents persisted up to 9 months after the primary vaccination series, as observed in studies that reported sex differences in antibody response among this population (29). This observation highlights the sustained impact of sex on vaccine-induced antibody production and suggests that the sex-based differences are not merely transient but may persist over an extended period after primary vaccination having implications on vaccine effectiveness and durability (37). Remarkably, sex-based differences in the persistence of antibody responses to influenza vaccination were associated with variations in the longevity of vaccine-induced immunity between males and females (38-40). Thus, this persistence of differences in the immune response to COVID-19 vaccination may have implications for the duration of protection against SARS-CoV-2, especially in high-risk populations such as NHRs.

Interestingly, these sex-based differences in antibody response were extinguished by booster doses, regardless of prior SARS-CoV-2 infection. This absence of sex differences in response to booster doses suggests that additional doses may effectively bridge potential immune response gaps between sexes and equalize immunity in both sexes (6,29,41). This leveling effect of the booster doses has also been reported among younger populations, where disparities in vaccine-induced antibody response due to age and sex, after the initial primary vaccination series, were found to be mitigated by a booster dose (42,43). Similarly, we did not detect differences between sexes to the Omicron BA.4/5 variant for the second monovalent booster and first bivalent booster doses.

There were similar rates of prior SARS-CoV-2 infection among both sexes across each vaccine dose. This suggests that prior exposure to SARS-CoV-2 did not significantly differ between males and females in this study. Therefore, the disparity in antibody response cannot be attributed to differences in previous infection rates. Also, we did not observe any significant difference in the time elapsed since COVID-19 infection between males and females among prior-infected residents. Thus, the sex-based differences in the antibody response are not attributable to differences in the timing of prior infection but are more likely influenced by inherent biological factors. For instance, in addition to the immunogenic effects of estrogen, the X chromosome contains immune-related genes, such as the Toll-like receptor 7, that can generate a more robust interferon (IFN) response, a critical component of the immune response to viral infections, including COVID-19 (44,45). With an enhanced IFN response, this X Chromosome-linked advantage contributes to a stronger and more effective immune reaction, including higher antibody production following vaccination.

It is essential to acknowledge the limitations of our study. Firstly, sex-specific comorbidities and frailty have been noted to influence antibody response among institutionalized older adults (29,30). Comorbidities and functional status were not collected for some of the early enrollees in our study. Among those with available comorbidity data, most male subjects were recruited from a state Veterans home. This presents both a distinct population and a different EHR for review than males in other community nursing homes. As comorbidities may be confounders that impact vaccine response (46), our finding at this timepoint still needs to be interpreted with caution. Secondly, we did not explore potential biological mechanisms that underlie the observed sex differences, such as the role of sex hormones and genetic factors. Also, we could not account for lifetime differences in prior exposure to respiratory viruses. Females have been reported to present more often with clinical viral respiratory infections than males (47) and therefore may have different lifetime immune priming, including, perhaps to other beta coronaviruses. Such a scenario could explain both an observation of less severity with SARS-CoV-2 infections (48) as well as differential initial boosting with the primary series, i.e., different anamnestic responses. Future research should delve deeper into these aspects to better understand the reasons behind these differences.

In conclusion, our study underscores the dynamic nature of sex-based differences in vaccine response and emphasizes the significance of booster doses in reducing these disparities. While a sex disparity was initially observed after the primary series, booster vaccinations effectively mitigated differences. These findings have implications for optimizing vaccine strategies in vulnerable populations and provide insight into the influence of prior infection on vaccine response in this specific context. Further research is required to unravel these sex-based differences in underlying mechanisms and implications and investigate their impact on COVID-19 outcomes.

## Data Availability

The de-identified dataset and related codes for analysis will be made available to researchers upon request after peer reviewed publication. Requests for data should be addressed to the corresponding authors.

## List of abbreviations

NHR: Nursing Home Resident
COVID-19: Coronavirus Disease 2019
WCG: Western-Copernicus Group
SARS-CoV-2: Severe Acute Respiratory Syndrome Coronavirus 2
mRNA: Messenger Ribonucleic Acid
RBD: Receptor Binding Domain
PCR: Polymerase Chain Reaction
BAU: Binding Antibody Unit
AU: Arbitrary Unit
LLD: Lower Limit of Detection
GMT: Geometric Mean Titer
CI: Confidence Interval

## STATEMENTS AND DECLARATIONS

### Authors’ contributions

*Concept and Design:* Oladayo A. Oyebanji, Sabra L. Klein, David H. Canaday, Brigid M. Wilson; *Manuscript preparation:* Oladayo A. Oyebanji, David H. Canaday, Brigid M. Wilson, Anna Yin, Patrick Shea; *Data analysis:* Brigid M. Wilson; *Data collection*: Oladayo A. Oyebanji, Alejandro B. Balazs, Jürgen Bosch, Christopher L. King, Nicholas Sundheimer, Vaishnavi Ragavapuram, Yi Cao; *Interpretation and funding:* Philip A. Chan, Aman Nanda, Rohit Tyagi, Sakeena Raza, Nadia Mujahid, Yasin Abul, Christopher L. King, David H. Canaday.

All authors reviewed the manuscript.

## Acknowledgments

Thank you to these individuals for their substantial assistance in various parts of the study.

*Case Western Reserve University:* Debbie Keresztesy, Olajide Olagunju, Dennis Wilk, Carson Smith, Alexandra Paxitzis, Htin Aung, Micheal Payne, Ellen See.

*Brown University & Lifespan:* Clare Nugent, Elizabeth White, Rosa Baier, Amy Recker, Joyce Sunday, Igor Vishnepolskiy, Evan Dickerson, Laurel Holland, Shreya Kamojjala, Alex Pralea, Tiffany Wallace, Lynn McNicoll.

